# Human origin ascertained for SARS-CoV-2 Omicron-like spike sequences detected in wastewater: a targeted surveillance study of a cryptic lineage in an urban sewershed

**DOI:** 10.1101/2022.10.28.22281553

**Authors:** Martin M. Shafer, Max J. Bobholz, William C. Vuyk, Devon Gregory, Adelaide Roguet, Luis A. Haddock Soto, Clayton Rushford, Kayley H. Janssen, Isla Emmen, Hunter J. Ries, Hannah E. Pilch, Paige A. Mullen, Rebecca B. Fahney, Wanting Wei, Matthew Lambert, Jeff Wenzel, Peter Halfmann, Yoshihiro Kawaoka, Nancy A. Wilson, Thomas C. Friedrich, Ian W. Pray, Ryan Westergaard, David H. O’Connor, Marc C. Johnson

## Abstract

**Background:** The origin of novel SARS-CoV-2 spike sequences found in wastewater, without corresponding detection in clinical specimens, remains unclear. We sought to determine the origin of one such “cryptic” wastewater lineage by tracking and characterizing its persistence and genomic evolution over time.

**Methods:** We first detected a cryptic lineage in Wisconsin municipal wastewater in January 2022. By systematically sampling wastewater from targeted sub-sewershed lines and maintenance holes using compositing autosamplers, we traced this lineage (labeled WI-CL-001) to its source at a single commercial building. There we detected WI-CL-001 at concentrations as high as 2.7 × 10^9^ genome copies per liter (gc/L) via RT-dPCR. In addition to using metagenomic 12s rRNA sequencing to determine the virus’s host species, we also sequenced SARS-CoV-2 spike receptor binding domains (RBDs), and where possible, whole viral genomes to identify and characterize the evolution of this lineage over the 13 consecutive months that it was detectable.

**Findings:** The vast majority of 12s rRNAs sequenced from wastewater leaving the identified source building were human. Additionally, we generated over 100 viral RBD and whole genome sequences from wastewater samples containing the cryptic lineage collected between January 2022 and January 2023. These sequences contained a combination of fixed nucleotide substitutions characteristic of Pango lineage B.1.234, which circulated in humans in Wisconsin at low levels from October 2020 to February 2021. Despite this, mutations in the spike gene, and elsewhere, resembled those subsequently found in Omicron variants.

**Interpretation:** We propose that prolonged detection of WI-CL-001 in wastewater represents persistent shedding of SARS-CoV-2 from a single human initially infected by an ancestral B.1.234 virus. The accumulation of convergent “Omicron-like” mutations in WI-CL-001’s ancestral B.1.234 genome likely reflects persistent infection and extensive within-host evolution.

**Funding:** The Rockefeller Foundation, Wisconsin Department of Health Services, Centers for Disease Control and Prevention (CDC), National Institute on Drug Abuse (NIDA), and the Center for Research on Influenza Pathogenesis and Transmission.

**Research in context:** *Evidence before this study:* To identify other studies that characterized unusual wastewater-specific SARS-CoV-2 lineages, we conducted a PubMed search using the keywords “cryptic SARS-CoV-2 lineages” or “novel SARS-CoV-2 lineages” in addition to “wastewater” on May 9, 2023. From the 18 papers retrieved, only two reported wastewater-specific cryptic lineages. These lineages were identified by members of our author team in wastewater from California, Missouri, and New York City. None of these could be definitively traced to a specific source. A third study in Nevada identified a unique recombinant variant (designated Pango lineage XL) in wastewater, which was also discovered in two clinical specimens from the same community. However, it was unclear whether the clinical specimens collected were from the same individual(s) responsible for the virus detected in the wastewater. To our knowledge, no prior study has successfully traced novel SARS-CoV-2 lineages detected in wastewater back to a specific location. How and where cryptic lineages are introduced into wastewater is not known.

*The added value of this study:* This study documents the presence and likely source of a novel and highly divergent cryptic SARS-CoV-2 lineage detected in Wisconsin wastewater for 13 months. In contrast to previously reported cryptic lineages, we successfully traced the lineage (WI-CL-001) to a single commercial building with approximately 30 employees. The exceptionally high viral RNA concentrations at the source building facilitated the tracing effort and allowed for the sequencing of WI-CL-001’s whole genome, expanding our view of the lineage’s mutational landscape beyond the spike gene.

*Implications of all the available evidence:* WI-CL-001’s persistence in wastewater, its heavily mutated Omicron-like genotype, and its identified point source at a human-occupied commercial building all support the hypothesis that cryptic wastewater lineages can arise from persistently infected humans. Because cryptic wastewater lineages have some amino acid changes that subsequently emerge in circulating viruses, increased global monitoring of such lineages could help forecast variants that may arise in the future.

## Introduction

SARS-CoV-2-infected hosts shed viral RNA in their stool and urine. Furthermore, the virus is known to infect the gastrointestinal (GI) tract and kidney tissues.^1,2^ Accordingly, SARS-CoV-2 RNA is readily detected in wastewater samples.^3^ Wastewater surveillance has thus become an important complement to clinical testing in monitoring SARS-CoV-2.^4^ Wastewater surveillance has also identified unique genetic lineages that clinical nasal swab testing has not.^5–6^ These SARS-CoV-2 lineages, which have been detected in wastewater but not in clinical specimens from individual persons, have been termed “cryptic” lineages. Although most cryptic lineages are not related to Omicron, they frequently have evolved mutational landscapes that are convergent with those of Omicron lineages.^6^

Compared to SARS-CoV-2 viruses that circulated in 2020, the Omicron lineage that was first detected in southern Africa in November 2021 (BA.1) had a highly divergent spike protein.^7^ The BA.1 spike protein had 12 lineage-defining amino acid substitutions in the receptor binding domain (RBD) between residues 412 and 579 (K417N, N440K, G446S, S477N, T478K, E484A, Q493R, G496S, Q498R, N501Y, Y505H, and T547K).^7–9^ Although global circulation of BA.1 did not begin until late 2021, ten of these RBD polymorphisms were observed in various combinations in sequence reads from New York City wastewater samples collected in the first half of 2021. One wastewater sample, collected in May 2021, had sequences encoding 24 amino acid substitutions in the spike RBD. Additional cryptic lineages were detected in wastewater samples from Missouri and California.^6^ It is not clear how or where these divergent SARS-CoV-2 sequences were introduced into wastewater.

There are two leading hypotheses for the source of these cryptic lineage sequences. First, an un- or under-sampled animal reservoir may be introducing these viruses into wastewater, e.g. through defecation in combined sewer systems, via surface and groundwater inflow/infiltration, or from livestock processing waste. SARS-CoV-2 exhibits a broad host range, including wild animals, livestock, and household pets.^10–12^ Multiple studies have found evidence of extensive SARS-CoV-2 transmission among wild white-tailed deer and farmed mink, initiated by infection from humans.^11–12^ There have also been examples of spillback into human hosts from each of these species.^13–14^ Thus, it is plausible that wild animal reservoirs of cryptic lineages exist undetected, with ongoing virus transmission and exchange between animal species.^15^

Alternately, cryptic SARS-CoV-2 lineage sequences in wastewater could be derived from people with unsampled infections.^16^ In a recent cohort study, 49.2% of participants had viral RNA (vRNA) in stool in the first week of infection; vRNA remained detectable in stool four months later in 12.7% of individuals even though all had cleared vRNA from the nasopharynx.^17^ Such individuals could contribute vRNA to wastewater even while testing negative via nasal swabs. Persons with immunocompromising conditions are at high risk for prolonged infections, and suboptimal immune responses in such individuals could select for antigenic variation over the course of infection, driving diversification of SARS-CoV-2 within these hosts.^18,19^ Such selection could account for the observation that cryptic lineages tend to accumulate high levels of nonsynonymous variation in spike while otherwise maintaining the characteristic mutations from viruses that are no longer common in circulation. In this study we identified a cryptic lineage (WI-CL-001) that was detectable at high levels in wastewater, documented its persistence and evolution over a one year period, pinpointed where it entered the wastewater stream and determined its human origin through a detailed sampling of the sewerage system.

## Methods

### Ethics statement

This activity was reviewed by CDC and the Wisconsin Department of Health Services and was conducted consistent with applicable federal law and CDC policy (see, e.g., 45 C.F.R. part 46, 21 C.F.R. part 56; 42 U.S.C. §241(d); 5 U.S.C. §552a; 44 U.S.C. §3501 et seq). The city, sewer plant, and locations of individual sampling sites have been concealed to protect the privacy of participants and residents. The authors representing the state public health authority had numerous conversations with the leadership of the identified source building in person and by phone, to explain the rationale for the investigation and describe the ways we would protect the identity of the company and employees. Specifically, we intentionally did not describe the investigation as a response to a known public health hazard, but rather as an activity to learn more about new technologies, and make sense of an unexpected finding that may someday be of public health significance. The wastewater findings were taken as a signal that people who work in the building may benefit from testing and linkage to care; once this testing was made available, we made it clear that the results would have the same privacy and confidentiality protections as all other testing.

### Sample collection

Wastewater samples for this study were collected from one metropolitan area in Wisconsin by wastewater engineers from the city wastewater utility. All wastewater samples were integrated over a 24-hour period using compositing ISCO 6712 and 6712c autosamplers (ISCO, Lincoln, NE, USA). The Wisconsin State Laboratory of Hygiene (WSLH), in consultation with utility engineers and the Wisconsin Department of Health Services (WDHS), determined specific testing locations in the waste-water collection system, allowing us to gradually narrow down the origin of WI-CL-001. Samples used for this investigation included daily wastewater samples from the central, publicly-owned treatment works (POTW) and twice-weekly monitoring of the primary sub-district lines (January 2022 through March 2023), which were collected through the Wisconsin Wastewater Monitoring Program. Additional sampling for this investigation included targeted maintenance hole testing from March 2022 through May 2022, directed at isolating the source of the cryptic lineage within the wastewater collection system. Further targeted sampling occurred at a commercial building sewer line access point (Facility Line B) serving six toilets on June 16^th^, August 16^th^, September 21^st^ and September 27^th^, 2022. Using this strategy, the sampled human source population of the WI-CL-001 signal was narrowed from more than 100,000 people to less than 30 people (Figure 1a). After consulting with local public health officials and facility managers, employees present at the facility were offered RT-PCR testing for SARS-CoV-2 testing via nasal swabs; 19 out of approximately 30 employees provided samples. Further methodological information about wastewater and clinical sample collection can be found in the appendix (pp. 2, 4).

**Figure 1.**
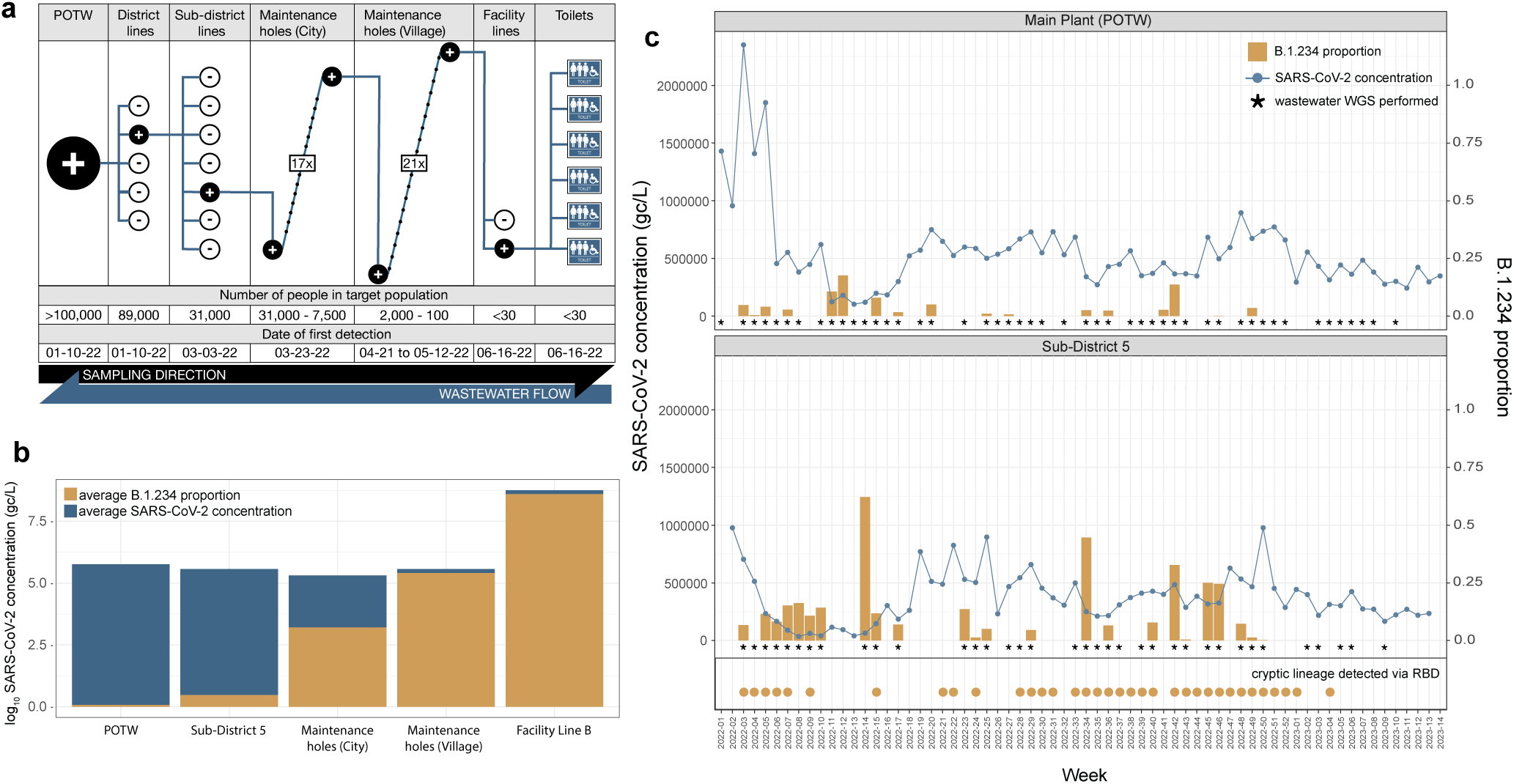
Tracing the source of the cryptic SARS-CoV-2 lineage. (a) WI-CL-001 was first detected at the publicly owned treatment works (POTW) facility from one of the five district lines that serve the POTW sewershed. This POTW serves a population of greater than 100,000 people. Continued wastewater sampling at sub-district lines that serve the positive district line isolated the lineage’s source to a single sub-district (Sub-District 5). Further sampling of maintenance holes within Sub-District 5 identified a single place of business as WI-CL-001’s source. Sampling at the commercial building facility pinpointed a collecting line (Facility Line B) servicing 6 toilets used by fewer than 30 facility employees. All location names are pseudonyms and all population values are approximate to conceal the identity of the sampled sewershed. (b) Average SARS-CoV-2 concentrations (on a log_10_ scale) obtained by RT-dPCR detected in five sampling areas show extremely high levels of SARS-CoV-2 in wastewater from Facility Line B. WI-CL-001’s contribution (B.1.234 proportion as estimated by Freyja within WGS data) to the SARS-CoV-2 concentration levels at each sampling level is shown in tan. (c) SARS-CoV-2 RT-dPCR concentrations throughout 2022 for the Main Plant (POTW) and Sub-District 5 are shown as a blue line. The percent contribution of WI-CL-001 (B.1.234 proportion in WGS data) is shown as tan bars, depicting the continued detection of the cryptic virus at both sampling levels for most of 2022. Higher B.1.234 proportions were seen in Sub-District 5 than in the Main Plant (POTW), corresponding to its closer proximity to WI-CL-001’s source. Asterisks mark the weeks in which samples were whole-genome sequenced, regardless of whether or not WI-CL-001 was detected in those sequences. Dates WI-CL-001 was detected in Sub-District 5 wastewater samples via targeted RBD amplicon sequencing are marked by tan circles. Weeks without tan circles were either negative or not tested via RBD sequencing.

### Procedures

Wastewater samples were shared between the Wisconsin State Laboratory of Hygiene (WSLH) and the University of Missouri, with the WSLH focusing on virus quantitation and whole genome sequencing, and the University of Missouri focusing on RBD-targeted sequencing. These two institutions had different procedures for viral RNA isolation, quantification, and sequencing.

At the WSLH, after the addition of a bovine coronavirus (BCoV) viral recovery control and the concentration of virus using Nanotrap Magnetic Virus Particles (Ceres Nanosciences, Manassas, VA, USA) on a Kingfisher Apex instrument (ThermoFisher Scientific, Waltham, MA, USA), total nucleic acids were extracted using Maxwell HT Environmental TNA kits (Promega, Madison, WI, USA) on a Kingfisher Flex instrument (ThermoFisher Scientific, Waltham, MA, USA).

The WSLH quantified the concentration of SARS-CoV-2, BCoV (viral recovery control), and PMMoV (fecal marker) in each sample using reverse transcriptase (RT) digital PCR (RT-dPCR). PCR inhibition was probed with a bovine respiratory syncytial virus (BRSV) spiked into each PCR reaction. Further details on our viral RNA isolation and quantification procedures are provided in the appendix (pp. 2-3).

For SARS-CoV-2 whole genome sequencing (WGS) at the WSLH, 13 µL of total nucleic acids from the WSLH’s wastewater extracts were used as input to Qiagen’s Direct SARS-CoV-2 Enhancer kit (Qiagen, Germantown, MD, USA). Amplicon libraries were prepared on a Biomek i5 liquid handler (Beckman Coulter, Brea, CA, USA). Libraries were quantified using a High Sensitivity Qubit 1X dsDNA HS Assay Kit (ThermoFisher Scientific, Waltham, MA, USA), and fragment size was analyzed by a QIAxcel Advanced and the QX DNA Screening Kit (Qiagen, Germantown, MD, USA). Sequencing was performed on an Illumina MiSeq instrument using MiSeq Reagent v2 (300 cycles) kits (Illumina, San Diego, CA, USA).

Fastq files were analyzed with the nf-core/viralrecon 2.5 workflow (<u>10.5281/zenodo.3901628</u>) using the SARS-CoV-2 Wuhan-Hu-1 reference genome (Genbank accession MN908947.3).^20^ The workflow was initiated as outlined on the project’s data portal (https://go.wisc.edu/4134pl). Additional details are provided in the appendix (pp. 3-4).

The University of Missouri concentrated the virus using a PEG protocol on pre-filtered wastewater samples (0.22 μM polyethersulfone membrane (Millipore, Burlington, MA, USA)). Samples were incubated with PEG (polyethylene glycol 8000) and 1.2 M NaCl, centrifuged, and the RNA was isolated from the pellet with the QIAamp Viral RNA Mini Kit (Qiagen, Germantown, MD, USA). Further information on these procedures can be found in the appendix (p. 2).

A nested RT-PCR approach was used to selectively amplify non-Omicron spike protein RBD regions from wastewater samples. Amplified RBD regions were then sequenced using an Illumina MiSeq instrument (Illumina, San Diego, CA, USA) and analyzed using the SAMRefiner software.^21^ WI-CL-001’s unique RBD sequences were used to identify and track the lineage across time and space. Additional details are provided in the appendix (p. 3).

The University of Missouri also conducted 12s rRNA sequencing to assess what species were contributing to Facility Line B in June of 2022. Additional details are provided in the appendix (p. 4)

### Statistical analyses

Variant proportions were assessed from WGS data using Freyja v.1.3.11, a tool previously developed to estimate the proportions of SARS-CoV-2 variants in deep sequence data containing mixed populations (<u>10.1038/s41586-022-05049-6</u>).^22^ Additional details are provided in the appendix (p. 5).

To generate the root-to-tip regression in Figure 4a, we used iqtree (v.2.2.0.3) to infer a maximum likelihood phylogenetic tree of all full SARS-CoV-2 consensus genomes belonging to Pango lineage B.1.234 (the inferred parent of WI-CL-001) from specimens collected in a 12-state Midwest region available in Genbank. Molecular clock rates and genetic distances were obtained through treetime (v0.9.3). Additional details and citations are provided in the appendix (p. 5). See Data Availability for accession numbers and scripts.

To assess the selective environment in which WI-CL-001 evolved, variant calls against reference genome MN908947.3 obtained through the nf-core/viralrecon workflows were processed using custom Python scripts to generate panels b and c in Figure 4. For panel b, variants were classified using SnpEff (v.5.0). For panel c, the proportion of nonsynonymous variants per site was calculated using SNPGenie (v.2019.10.31), and a binomial probability distribution was implemented using SciPy’s binomtest function (v.1.9.3). A Mann-Whitney two-sided test was applied to test the difference between πN and πS in each gene, while a one-sided test was used to test for an enrichment of the πN value of spike against the πN value on the other genes. For panel d the average Hamming distance between B.1.234 isolates (dataset used in Figure 4a) and the MN908947.3 reference sequence was calculated to obtain synonymous and nonsynonymous divergence values. Additional details and citations are provided in the appendix (p. 5). See Data Availability for accession numbers and scripts.

### Role of the funding source

This manuscript underwent CDC’s clearance review process due to the involvement of CDC co-authors. However, the funders did not play a direct role in the study design, data collection, data analysis, or manuscript preparation.

## Results

On January 11, 2022, a cryptic lineage containing at least six unusual spike RBD substitutions (V445A, Y449H, N460K, E484Q, F490Y, Q493K) was first detected in a composite wastewater sample from a metropolitan publicly owned treatment works (POTW) in Wisconsin (Figure 1c). Over the following six months, the source of the enigmatic RBD sequences was narrowed to a single commercial building, and subsequently a single sewer line serving six toilets (called “Facility Line B”) (Figure 1a). As the sampling effort progressively approached the source, the proportion of WI-CL-001 (labeled B.1.234 in Figure 1) increased relative to the total SARS-CoV-2 sequences detected at each sampling site (Figure 1b, 1c).

Consistently high wastewater SARS-CoV-2 RNA concentrations were observed in all samples collected from Facility Line B. Concentrations of ~5.2 × 10^8^, ~1.6 × 10^9^, ~2.7 × 10^9^, and ~5.5 × 10^8^ genome copies per liter (gc/L) undiluted wastewater were quantified using RT-dPCR on samples collected June 16th, August 16th, September 23rd, and September 27th, respectively (Figure 1b). Despite these high RNA concentrations, viable virus could not be cultured from wastewater after multiple attempts (appendix p. 4). 12s rRNA sequencing detected predominantly human rRNA from this source. Chicken rRNA, the next largest taxon identified, was less than 0.05% of the sample (appendix p. 9). Despite these findings, none of the building occupants who volunteered for nasal swab testing organized by the local public health department were positive for SARS-CoV-2 in June 2022. The wastewater signal from the Sub-District 5 line became undetectable in January 2023 after a decline in signal concentration beginning the 46th week of 2022 (Figure 1c).

The high levels of viral RNA in Facility Line B facilitated the amplification and sequencing of entire viral genomes to shed further light on the origins and evolution of this unusual lineage. We generated whole genome sequences from the Facility Line B samples taken on June 16th, August 16th, September 23rd, and September 27th, 2022. At the consensus level, all of these sequences were classified as lineage B.1.234 by Pangolin. In SARS-CoV-2 genomic surveillance using clinical specimens, B.1.234 viruses were detected in Wisconsin between September 2nd, 2020, and March 30th, 2021.^23^

The B.1.234 lineage does not have any characteristic spike RBD amino acid changes relative to the reference Wuhan-Hu-1 (Figure 2a). Sequencing single amplicons spanning the RBD allowed us to define haplotypes, i.e., specific combinations of mutations found together in a single RNA molecule. We repeatedly sequenced spike RBD in wastewater samples from the Sub-District 5 line and haplotypes of WI-CL-001 were detected every month from January 2022 to January 2023 (appendix p. 7). In all, we detected 54 RBD haplotypes between Jan 2022 and Jan 2023, with an average of 1.35 haplotypes (range, 1-3) being detected at any one timepoint (appendix p. 7). Some of the amino acid substitutions on these haplotypes predated the emergence of the same amino acid changes, or different changes at the same amino acid residues, in globally circulating Omicron lineages (Figure 3).^24^

**Figure 2.**
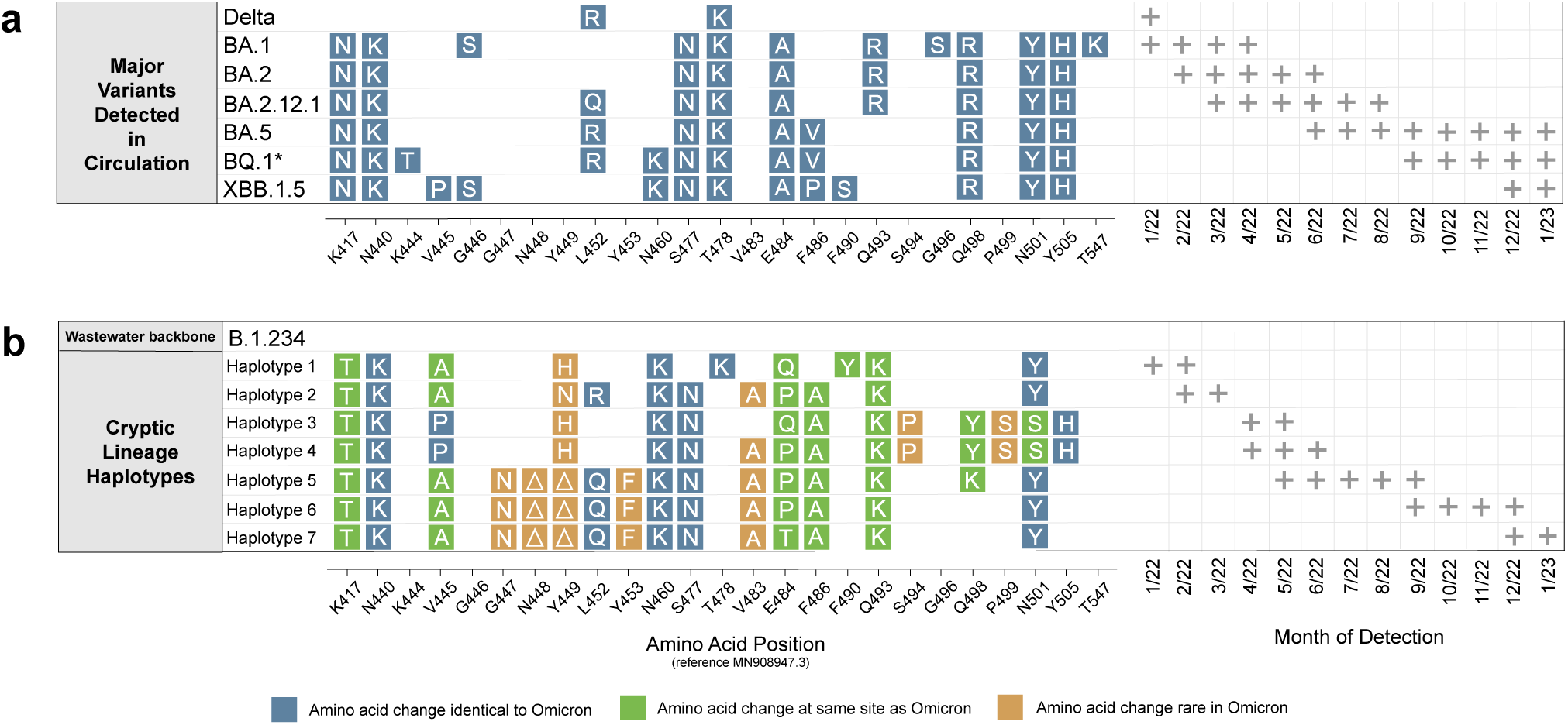
Representative haplotypes of WI-CL-001 sequences. (a) Delta and Omicron amino acid changes (shown in blue) represent characteristic changes seen in at least 90% of all US sequences of the specified sublineage.^23^ Plus signs on the right denote months in which these lineages were identified in at least 1% of all US clinical sequences. (b) Representative haplotypes detected from Sub-District 5 using non-Omicron spike RBD sequencing are displayed, each of which were at least 25% of the total reads in at least one sample. Green boxes indicate amino acid sites that are also altered in major Omicron lineages, blue boxes indicate amino acid sites that have mutations identical to major Omicron lineages, and tan boxes indicate amino acid sites that are altered in WI-CL-001 yet altered in less than 0.1% of Omicron sequences.^24^ Plus signs on the right indicate months in which these haplo-types were detected.

**Figure 3.**
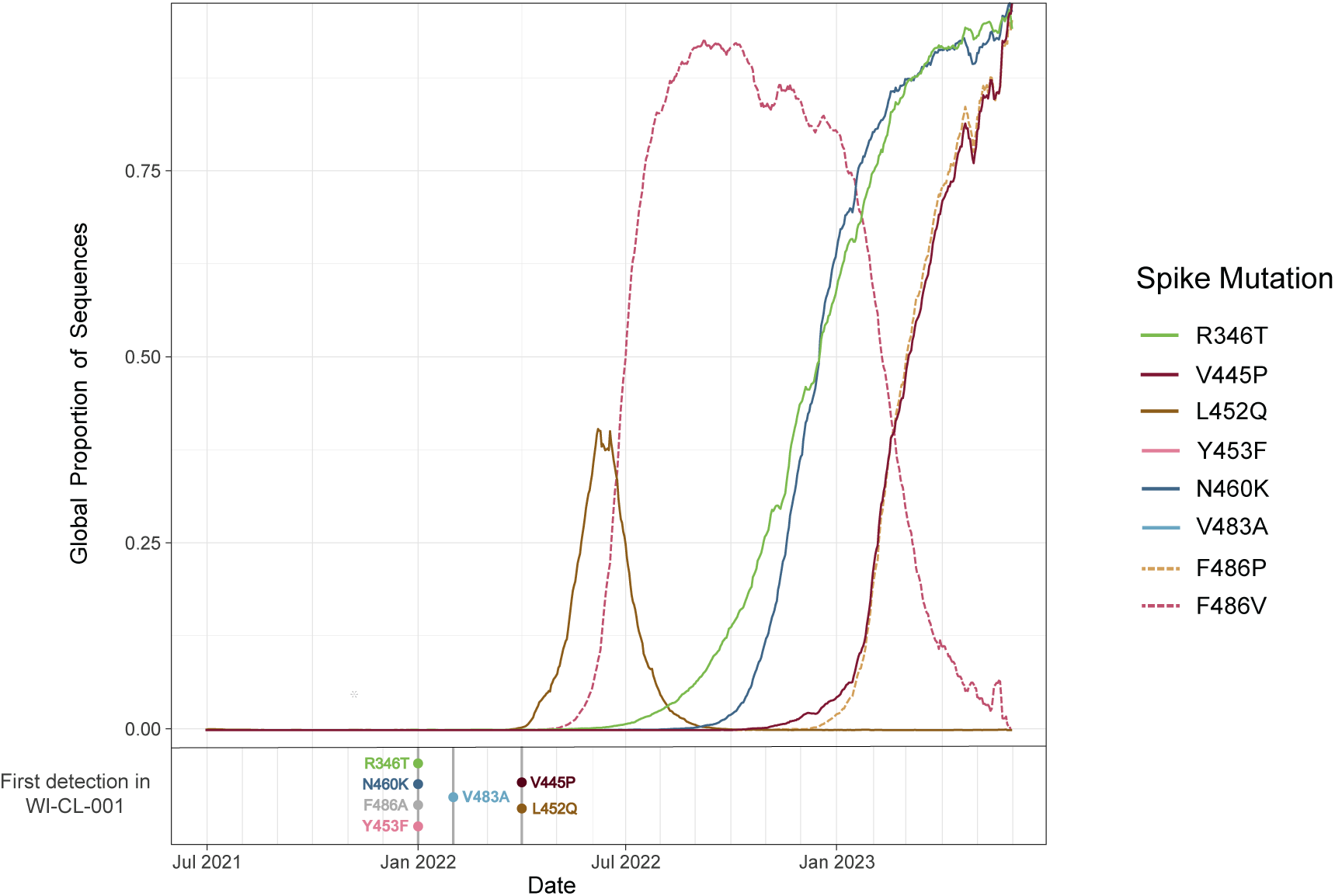
Prevalence of key cryptic lineage mutations in global sequences. The global proportions of sequences uploaded to NCBI GenBank between June 31st 2021 and May 1st 2023 for key mutations in the spike gene of the Wisconsin wastewater lineage are plotted over time.^23^ The spike mutations R346T, V445P, and N460K were all detected in the Wisconsin cryptic lineage months before becoming predominant in global sequences. WI-CL-001 also harbored F486A from the time of initial detection in January 2022. Two other substitutions at spike amino acid residue 486 have since become dominant in global sequences (dotted lines).

The cryptic lineage is also divergent outside of the spike RBD. When plotted on a radial phylogenetic tree using Nextclade, Illumina whole genome consensus sequences from Facility Line B show divergence from the Wuhan-Hu-1 reference similar to clade 22B and XBB* Omicron lineages (appendix p. 8).^25^ To investigate this further, iVar output from the nf-core/viralrecon workflow identified variants at ≥25% frequency (appendix p. 10, appendix 2).^20^ One of these mutations is in the N-terminal ectodomain of the membrane protein, where a 15 nucleotide in-frame insertion (I8delinsSNNSEF) is present at an average frequency of 92.4% in all Facility Line B whole genome sequences (appendix pp. 11-13).

We next asked whether the unusual combinations of mutations present in WI-CL-001 could be the result of natural selection favoring nonsynonymous (amino-acid-changing) mutations. First, we found that mutations accumulated in WI-CL-001 faster than expected based on the nucleotide substitution rate that prevailed when B.1.234 viruses were circulating in the US Midwest (Figure 4a). Across the four timepoints with available genome sequences, there is a notable excess of nonsynonymous nucleotide substitutions (mean 121.8 ± 16.3) relative to synonymous ones (mean 22.5 ± 4.7) (Figure 4b). To further characterize genetic diversity within each sample, we used the summary statistic π, which quantifies the number of pairwise differences per nonsynonymous (πN) and synonymous (πS) site within a set of sequences. Within the spike gene, πN was significantly greater than πS at each time-point, which could indicate ongoing diversifying selection on spike (Figure 4c). Spike also had significantly higher nonsynonymous diversity compared to ORF1ab, ORF3a, M, ORF6, ORF7a, and N at each timepoint (Figure 4c). Because π counts pairwise differences per site within a sample, mutations that have become fixed or nearly fixed within the virus population do not contribute to π values. We therefore next calculated divergence, i.e., the average Hamming distance between each sequenced virus (either B.1.234 variants or WI-CL-001) and the ancestral sequence Wuhan-Hu-1 (MN908947.3; Figure 4d). Both synonymous and nonsynonymous divergence values were substantially higher for WI-CL-001 than for B.1.234 viruses. Taken together, these findings suggest that WI-CL-001 sequences have been shaped by diversifying selection.

**Figure 4.**
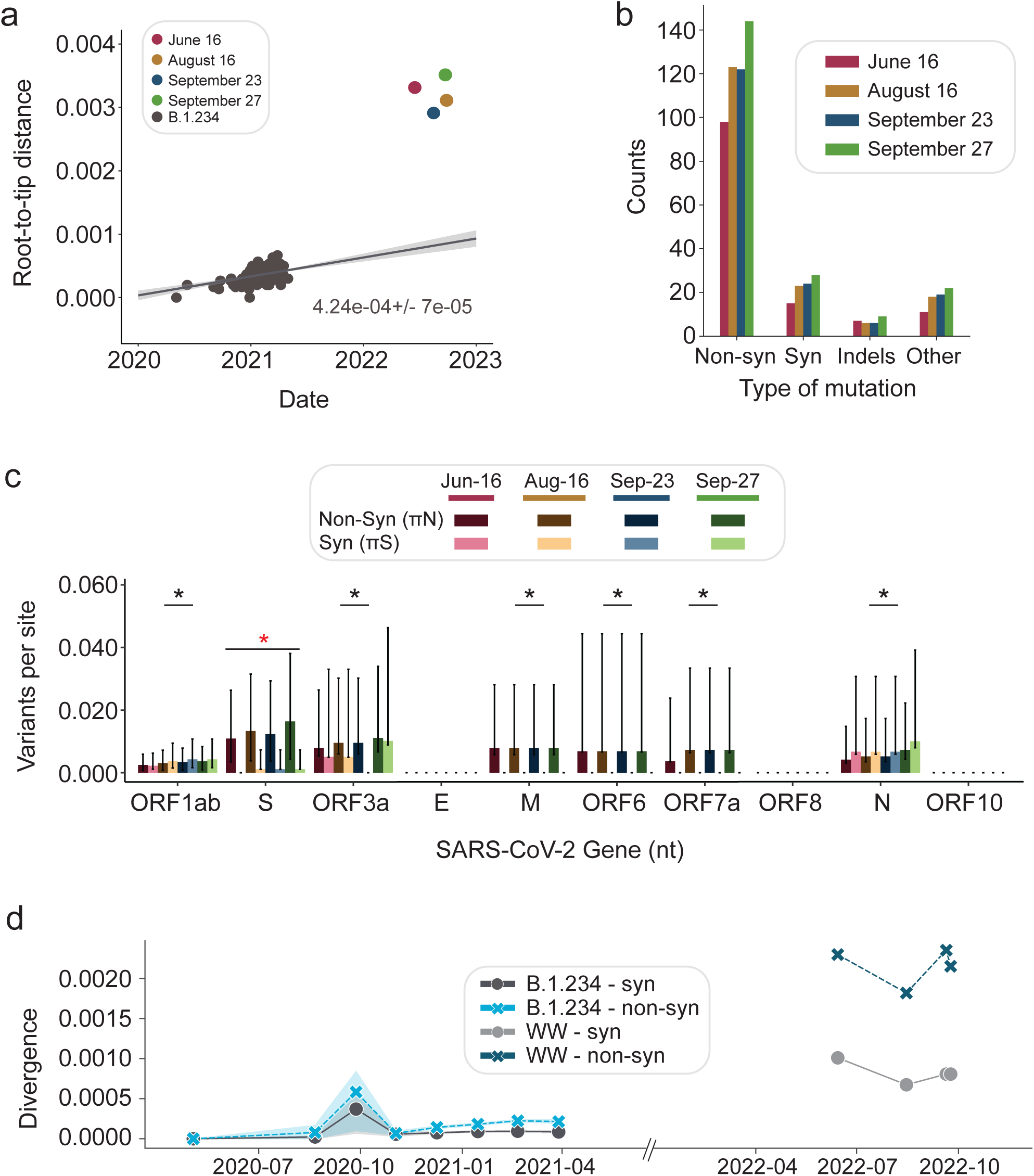
Analysis of wastewater genomic sequences from all Facility Line B timepoints. (a) Root-to-tip regression analysis (distance) of B.1.234 sequences via TreeTime based on a maximum likelihood phylogenetic tree inferred with iqtree (not shown) and aligned to Wuhan-Hu-1 (MN908947.3). All sequences were obtained from GenBank and can be accessed using the accession numbers available on the GitHub repository accompanying this manuscript. (b) Number of intra-sample single nucleotide polymorphisms (iSNVs; y-axis) for the wastewater timepoints for each mutation type relative to the reference MN908947.3 (colored as in panel a). Mutations were classified as nonsynonymous (Non-syn), synonymous (Syn), insertions-deletions (Indels), or others (including nonsense and frameshift mutations outside of coding regions). (c) Within-sample pairwise sequence diversity assessed for nonsynonymous sites (πN, darker bars) and synonymous sites (πS, lighter bars) in each SARS-CoV-2 gene at each timepoint. The 95% confidence intervals were obtained using a binomial probability distribution. A Mann-Whitney two-sided test was applied to test the difference between πN and πS on each gene (red asterisk). A one-sided test was used to test for an enrichment of the πN value of spike against the πN value of the other genes (black asterisks). (d) The divergence (Hamming distance; y-axis) between B.1.234 isolates from panel (**a**) and the MN908947.3 reference sequence over a sliding window of 36 days (x-axis) compared to WI-CL-001 isolates. Except for WI-CL-001, data are only plotted when windows contain at least two B.1.234 sequences.

## Discussion

We traced the source of a cryptic SARS-CoV-2 lineage, first detected in wastewater from a central municipal wastewater treatment facility, to a sewer line within a commercial building (Facility Line B). Non-human animal sequences made up a negligible proportion of 12s rRNA detected within Facility Line B, making an animal source highly improbable (appendix p. 9). The lineage was not detected by voluntary nasal swab testing at the source building, suggesting that a multi-person outbreak was not a likely source of the cryptic signal. Combining our observations, we posit that the simplest explanation for the appearance and persistence of WI-CL-001 is that a single human individual, originally infected when B.1.234 was in circulation, developed a persistent infection and continued to excrete viruses into wastewater throughout 2022. The WI-CL-001 signal became undetectable after 53 weeks, at the end of a multi-week decline in signal strength (Figure 1c). This is one of the longest periods of continuous detection that we are aware of for a SARS-CoV-2 cryptic lineage.^6^

The average SARS-CoV-2 concentration we detected in samples from Facility Line B (1.3 × 10^9^ gc/L) was 7 log2 fold change higher by RT-dPCR than the highest signal previously measured out of over 12,000 wastewater samples collected throughout Wisconsin between 2020 and 2023. Upon searching Pubmed for comparable results in other studies using the terms (((SARS-CoV-2) AND (wastewater)) AND (concentration)) AND (building) on September 15th 2023, none of the 40 papers we found reported a single-building wastewater SARS-CoV-2 concentration above 10^8^ gc/L. The high source concentration of WI-CL-001 may help to resolve a paradox from earlier cryptic lineage studies: if cryptic lineages come from only a single source, how could they be detected in a dilute municipal wastewater sample? Based on wastewater flow data from the Sub-District 5 line and estimations of typical toilet use, we would expect the WI-CL-001 viral RNA to be diluted from a wastewater volume of approximately 200 gallons at Facility Line B into a volume of 8 million gallons at the Sub-District 5 line. Thus, if there were 2 billion gc/L at Facility Line B, we would expect to detect ~50,000 gc/L at the Sub-District 5 line. This value is comparable to what we actually observed over 13 months (Figure 1c). Thus, our observations at both Facility Line B and Sub-District 5 are consistent with a persistently infected individual shedding very high amounts of SARS-CoV-2 RNA into wastewater at the source building over the course of 2022.

The large preponderance of nonsynonymous substitutions in the Facility Line B viral genomes suggests that this virus has undergone diversifying selection on spike, and perhaps on other genes. This is consistent with reports of individuals with prolonged SARS-CoV-2 infections, in whom weak immunity and persistent virus replication result in the selection of immune escape variants.^18,19^ Many RBD amino acid changes present in WI-CL-001 have eventually appeared in Omicron variants circulating in human populations. In the RBD region of the spike gene, R346T, V445P, L452Q, L452R, N460K, and F486V and F486P emerged in circulating Omicron variants globally between January of 2022 and January of 2023 (Figure 2). Some of these spike mutations, specifically R346T, V445P, and N460K emerged in WI-CL-001 five to six months before becoming highly prevalent globally, largely associated with the spread of BQ.1.1* and XBB.1.5. In WI-CL-001, a phenylalanine-to-alanine substitution at spike residue 486 (F486A) appeared approximately four months before the rise of S:F486V (found in BA.5*/BQ.1* variants) and ten months before the rise of S:F486P (found in XBB.1.5*). The RBD mutations Y453F and V483A were detected in WI-CL-001 respectively from January and February 2022 through January 2023 but were found in less than one percent of global sequences during that same time (Figure 3).^23^ We therefore speculate that those two substitutions, or other mutations at these sites, may become more prevalent in circulating viruses in the future. Since we made this prediction in May 2023, the mutation V483Δ has arisen at the same residue in BA.2.86.^26^

In addition to the highly divergent spike, there was a cluster of nearly-fixed variants in the region that encodes the ectodomain of the viral membrane protein. The mutation cluster includes a 15-nucleotide insertion (5’-GCAACAACTCAGAGT-3’) that encodes the amino acids SNNSEF by splitting the A and TT of an existing isoleucine codon. Interestingly, the insertion is identical to the sequence found between positions 11,893 and 11,907 in ORF1ab, which suggests an intramolecular recombination event. Additionally, WI-CL-001 has M:A2E, M:G6C, and M:L17V amino acid substitutions. The phenotypic impact of these substitutions is unclear. One possible explanation is that these substitutions confer escape from membrane protein-directed antibodies. This region of the membrane protein is exposed outside of the SARS-CoV-2 virion and is a known target for binding antibodies.^27,28^ In fact, a previous study using linear peptide binding arrays found that antibody binding on the membrane protein ectodomain was the most intense, on average, of all epitopes in the SARS-CoV-2 proteome.^28^ Together these observations suggest that the accumulation of mutations in WI-CL-001, particularly in spike (but in other genome regions as well), is the result of adaptive evolution.

WI-CL-001, BA.1, and the recently described BA.2.86 are all spike saltations, i.e., evolutionary “jumps”, involving the concurrent appearance of a large number of substitutions. These viruses share two common features: (1) in-frame insertions of multiple amino acids in regions frequently recognized by antibodies and (2) an extraordinary excess of non-synonymous substitutions in spike.^8,26^ BA.1 and BA.2.86 have insertions in the spike N-terminal domain that are reminiscent of the membrane protein ectodomain insertion observed in WI-CL-001. Indeed, the characteristic BA.2.86 insertion of ‘TCATGCCGCTGT’ is, like WI-CL-001’s membrane protein insertion, an apparent intramolecular recombination that is identical to a nucleotide sequence from ORF1ab (17166-17177). The striking excess of non-synonymous variants and near absence of synonymous variants in the spike protein for these three lineages suggests that they have each evolved under diversifying selection. While the original sources of BA.1 and BA.2.86 will never be known definitively, a leading hypothesis for the origin of some divergent SARS-CoV-2 variants is that they arise in immunocompromised individuals with prolonged infections.^18,19^

Accordingly, more frequent global wastewater viral surveillance/sequencing of catchment areas would likely detect many more examples of cryptic SARS-CoV-2 lineages. Traceback studies such as these, if conducted within appropriate ethical and privacy constraints, may be useful for determining the origins, as well as epidemiologic and clinical significance, of these lineages. We speculate that Omicron-derived cryptic lineages will be detectable in wastewater in the future. Given the extensive spread of Omicron, we expect the number of prolonged infections that give rise to these cryptic lineages to increase, making the emergence and detection of cryptic lineages more common. Although RBD sequencing covers only a small segment of the SARS-CoV-2 genome, we believe this method will continue to be valuable in wastewater surveillance due to its high sensitivity. Cryptic wastewater lineages like WI-CL-001 may or may not have the potential for wider community transmission. Nevertheless, the fact that these lineages frequently exhibit specific mutations, or changes at specific sites, that are later found in circulating variants could be used to aid in forecasting the future evolutionary trajectory of SARS-CoV-2. Such a forecast would be useful in evaluating the cross-protection of existing and future vaccines and treatments. In the present, wastewater sequencing surveillance has become a valuable approach for tracking the emergence of novel SARS-CoV-2 variants in the context of waning clinical sequencing and otherwise-unsampled prolonged infections.

## Data availability

Sequencing data are available in NCBI SRA and Genbank. Additional data is available from https://go.wisc.edu/4134pl. All sequences used for the phylogenetic inferences shown in Figure 4 were obtained from GenBank and can be accessed using the accession numbers available on the GitHub repository accompanying this manuscript, which also contains the scripts used to analyze them (https://github.com/tcflab/wisconsin_cryptic_lineages).

## Acknowledgments

This study was made possible by the generous support of the Rockefeller Foundation’s Regional Accelerators for Genomics Surveillance (DHO/TCF), Wisconsin Department of Health Services Epidemiology and Laboratory Capacity funds (www.dhs.wisconsin.gov, 144 AAJ8216) to DHO, CDC contract 75D30121C11060 (DHO/TCF), Wisconsin Department of Health Services ELC Wastewater Surveillance funds (www.dhs.wisconsin.gov, 130:AAI8627) to the UW-Madison Wisconsin State Laboratory of Hygiene (WSLH), and NIDA contract 1U01DA053893-01 (MJ), the Center for Research on Influenza Pathogenesis and Transmission (75N93021C00014) from the National Institutes of Allergy and Infectious Diseases to YK. The authors thank Roger Wiseman, Nick Minor, David Baker, and CDC SPHERES for helpful discussions. The authors also thank Sarah Abu Kamal, Maansi Bhasin, Sydney Wolf, and Aanya Virdi for help with sequence generation and data organization. They would also like to acknowledge and thank the wastewater engineers from the city wastewater utility for their sewershed sampling prowess. Additional thanks to Katia Koelle and Michael Martin of Emory University for helpful discussions on the quantitative analysis of viral evolution.

## Competing interests

YK has received unrelated funding support from Daiichi Sankyo Pharmaceutical, Toyama Chemical, Tauns Laboratories, Shionogi, Otsuka Pharmaceutical, KM Biologics, Kyoritsu Seiyaku, Shinya Corporation and Fuji Rebio.

## Notes

### Author Declarations

All human subjects work was conducted by the CDC and the Wisconsin Department of Health Services and was consistent with applicable federal law and CDC policy (see ethics statement).

### Summary of Updates

The manuscript has been revised to more clearly articulate the comparison between Omicron and the Wisconsin cryptic lineage (now labelled WI-CL-001). Summary, Methods, Results and Discussion text have been re-apportioned to better fit with Lancet Microbe guidelines. The in-text methods section is now more comprehensive than previous versions. More context is given in discussion of observed SARS-CoV-2 RNA concentrations in wastewater. Author Isla E Emmen added.

