## Appendix for "Human origin ascertained for SARS-CoV-2 Omicron-like spike sequences detected in wastewater: a targeted surveillance study of a cryptic lineage in an urban sewershed"

### Supplementary Methods

#### Collection of wastewater

Wastewater samples for this study (January 2022 through March 2023) were collected from one metropolitan area in Wisconsin by experienced wastewater engineers from the city wastewater utility. The Wisconsin State Laboratory of Hygiene (WSLH) and Wisconsin Department of Health Services (WDHS) determined specific locations in the wastewater collection system to obtain samples for each round of testing, allowing them to gradually narrow down the origin of the WI-CL-001 source region. Sewage lift-stations, maintenance holes, and facility sewer line access points were sampled with compositing ISCO 6712 and 6712c autosamplers (ISCO, Lincoln, NE, USA). Depending upon maintenance hole depth, the autosampler was either placed on a shelf adjacent to the wastestream or suspended from the maintenance hole opening, with weighted collection lines placed into the wastewater stream. The city wastewater utility provided the estimated population size served for each collection point along the sewer line. All autosamplers were programmed to collect 24-hr composites, typically on a time-based mode, with wastewater composited into a 10-liter polypropylene container. The composite was kept cool during collection with ice packed around the collection container. Composite samples were transported to the analytical laboratory within a few hours of sample retrieval. While wastewater flows were available from the pump-stations and central municipal wastewater treatment facility, flow measurements were not made in the maintenance hole waste streams.

#### Isolation of viral RNA from wastewater

Two approaches were used to isolate viral RNA from wastewater.

For samples processed at WSLH, wastewater samples (homogenized and unfiltered) were spiked with 20 µL/250 mL Calf-Guard® (Zoetis, Parsippany, NJ, USA), a cattle vaccine containing Bovine Coronavirus (BCoV) (as a virus recovery control), and briefly stored at 4°C until the viral targets were isolated and concentrated, typically on the day of receipt. A total of 10 mL (2x5mL) of wastewater was concentrated using Nanotrap Magnetic Virus Particles, Microbiome A and Enhancing Reagent 2 (Ceres Nanosciences, Manassas, VA, USA), using a KingFisher Apex automation platform (ThermoFisher Scientific, Waltham, MA, USA). Total nucleic acids (TNA) were extracted using Maxwell(R) HT Environmental TNA kits (Promega, Madison, WI, USA) and eluted in 200 µL of 25 mM Tris HCl (pH 8.0) buffer. The extraction was automated using a KingFisher Flex (ThermoFisher Scientific, Waltham, MA, USA). KingFisher programs are available on Figshare: https://doi.org/10.6084/m9.figshare.21538143.v4.The "expanded"program was used for the concentration. A method blank made of 1X PBS spiked with BCoV was processed for each concentration/extraction batch along with an extraction blank (sample replaced by sterile water).

For samples processed at the University of Missouri, samples were processed as previously described.[1] Briefly, wastewater samples were centrifuged at 3000×g for 10 min and filtered through a 0.22 µM polyethersulfone membrane (Millipore, Burlington, MA, USA). Approximately 37.5 mL of wastewater was mixed with 12.5 mL solution containing 50% (w/vol) polyethylene glycol 8000 and 1.2 M NaCl, mixed, and incubated at 4C for at least 1 h. Samples were then centrifuged at 12,000×g for 2 h at 4C. Supernatant was decanted and RNA was extracted from the remaining pellet (usually not visible) with the QIAamp Viral RNA Mini Kit (Qiagen, Germantown, MD, USA) using the manufacturer's instructions. RNA was extracted in a final volume of 60 µL.

#### Quantification of viral RNA by RT-dPCR

Quantification of SARS-CoV-2, BCoV (internal control), PMMoV (fecal marker), and BRSV (RT-PCR spiked-in inhibition control) was achieved using reverse transcriptase digital PCR (RT-dPCR). Master mix was prepared using the One-Step Viral PCR kit (4x) (Qiagen, Germantown, MD, USA) and GT dPCR SARS-CoV-2 Wastewater Surveillance Assay Kit (GT Molecular, Fort Collins, CO, USA) with quantification of the following viral targets: N1, N2, BCoV, and PMMoV included with the GTMolecular dPCR SARS-CoV-2 Wastewater Surveillance Assay Kit, and BRSV primers and probes from IDT.[2] The samples were quantified on a QIAcuity Four Digital PCR System (Qiagen, Germantown, MD, USA). N1, N2, and BCoV were multiplexed on QIAcuity Nanoplate 26k 24-well plates while PMMoV and BRSV were singleplexed on 8.5k 96-well nanoplates. Cycling and exposure conditions are detailed in the table shown below. Analysis of the RT-dPCR results was performed with the QIAcuity Software Suite version 2.1.7.182. Thresholds were manually set to separate negative and positive partitions. For all targets and RT-dPCR runs, a non-template control and the method and extraction blanks were tested to assess any contamination. Positive controls were tested to ensure the RT-PCR setup and reagent integrity. When inhibition was detected, samples were tested at an higher dilution.

**Table. dPCR Thermocycling Conditions:**

| Thermocycling Conditions: | | | |
|---|---|---|---|
| **Step** | | **Time** | **Temp °C** |
| Reverse Transcription | | 30 min | 50 |
| DNA polymerase activation | | 2 min | 95 |
| **45 cycles** | Denaturation | 10 sec | 95 |
| | Anneal/Extend | 30 sec | 55 |
| Target | Channel | Exposure | Gain |
| N1 | Red (ROX) | 500 | 4 |
| N2 | Green (FAM) | 300 | 6 |
| BCoV | Yellow (HEX) | 300 | 6 |
| PMMoV | Green (FAM) | 300 | 6 |
| BRSV | Yellow (HEX) | 500 | 6 |

**Identification of cryptic lineages in wastewater with non-Omicron RT-PCR amplification and amplicon sequencing**

The primary RBD RT-PCR was performed using the Superscript IV One-Step RT-PCR System (Thermo Fisher Scientific,12594100, Waltham, MA, USA). Primary RT-PCR amplification was performed as follows: 25 °C (2:00) + 50 °C (20:00) + 95 °C (2:00) + [95 °C (0:15) + 55 °C (0:30) + 72 °C (1:00)] × 25 cycles using the MiSeq primary PCR primers 5'-ATTCTGTCCTATATAATTCCGCAT-3' and 5'-CCCTGATAAAGAACAGCAACCT-3' (the first primer was changed to 5'-TATATAATTCCGCATCATTTTCCAC-3' starting in May, 2022 to adapt to changing Omicron lineages). Secondary PCR (25 µL) was performed on RBD amplifications using 5 µL of the primary PCR as template with MiSeq nested gene specific primers containing 5′ adapter sequences (0.5 µM each) 5'-acactctttccctacacgacgctcttccgatctGTGATGAAGTCAGACAAATC-GC-3' and 5'-gtgactggagttcagacgtgtgctcttccgatctATGTCAAGAATCTCAAGTGTCTG-3', dNTPs (100 µM each) (New England Biolabs, N0447L) and Q5 DNA polymerase (New England Biolabs, M0541S, Ipswich, MA, USA). Secondary PCR amplification was performed as follows: 95 °C (2:00) + [95 °C (0:15) + 55 °C (0:30) + 72 °C (1:00)] × 20 cycles. A tertiary PCR (50 µL) was performed to add adapter sequences required for Illumina cluster generation with forward and reverse primers (0.2 µM each), dNTPs (200 µM each) (New England Biolabs, N0447L, Ipswich, MA, USA) and Phusion High-Fidelity or (KAPA HiFi for CA samples) DNA Polymerase (1U) (New England Biolabs, M0530L, Ipswich, MA, USA). PCR amplification was performed as follows: 98 °C (3:00) + [98 °C (0:15) + 50 °C (0:30) + 72 °C (0:30)] × 7 cycles +72 °C (7:00). Amplified product (10 µl) from each PCR reaction is combined and thoroughly mixed to make a single pool. Pooled amplicons were purified by the addition of Axygen AxyPrep MagPCR Clean-up beads (Corning, MAG-PCR-CL-50, Corning, NY, USA) or in a 1.0 ratio to purify final amplicons. The final amplicon library pool was evaluated using the Agilent Fragment Analyzer automated electrophoresis system (Agilent, Santa Clara, CA, USA), quantified using the Qubit HS dsDNA assay (ThermoFisher Scientific, Waltham, MA, USA), and diluted according to Illumina's standard protocol. The Illumina MiSeq instrument was used to generate paired-end 300 base pair reads (Illumina, San Diego, CA, USA). Adapter sequences were trimmed from output sequences using Cutadapt.

Sequencing reads were processed as previously described. Briefly, VSEARCH tools were used to merge paired reads and dereplicate sequences.[3] Dereplicated sequences from RBD amplicons were mapped to the reference sequence of SARS-CoV-2 (NC_045512.2) spike ORF using Minimap2.[4] Mapped amplicon sequences were then processed with SAM Refiner using the same spike sequence as a reference and the command line parameters "--Alpha 1.8 --foldab 0.6".[5]

The haplotypes representing at least 25% of the total sequences in at least one sample were rendered into figures using plotnine (https://plotnine.readthedocs.io/en/stable/index.html).

**SARS-CoV-2 whole genome sequencing of wastewater**

Sequencing libraries were generated at the WSLH using the QIAseq DIRECT SARS-CoV-2 Enhanced kits with the primer Booster (Qiagen, Germantown, MD, USA) following manufacturer's instructions. Briefly, 13 uL of total nucleic acid were reverse transcribed into cDNA using hexaprimers. SARS-CoV-2 genome was then specifically enriched using a SARS-CoV-2 primer panel. The panel consists of approximately 550 primers for creating 425 amplicons, covering the entire SARS-CoV-2 viral genome. UDI were 1:5 diluted. The library preparation was fully automated using the Biomek i5 Automated

Workstation (Beckman Coulter, Brea, CA, USA). Libraries were quantified using a High Sensitivity Qubit 1X dsDNA HS Assay Kit (ThermoFisher Scientific, Waltham, MA, USA) and fragment size analyzed by a QIAxcel Advanced and the QX DNA Screening Kit (QIAGEN, Germantown, MD, USA). Libraries were sequenced on an Illumina MiSeq platform using MiSeq Reagent v2 (300 cycles) kits (Illumina, San Diego, CA, USA).
Isolated RNA from each Facility Line B time point was whole-genome sequenced at least twice in separate Illumina MiSeq runs in anticipation of needing sequence technical replicates for later analysis. The data were analyzed with the nf-core/viral-recon workflow (https://nf-co.re/viralrecon/2.5) using the SARS-CoV-2 Wuhan-Hu-1 reference genome (Genbank accession MN908947.3) and the QIAseq Direct SARS-CoV-2 primer .bed file (https://www.qiagen.com/us/products/next-generation-sequencing/rna-sequencing/qiaseq-direct-sars-cov-2-kits/).[6] After creating a sample sheet as described on the nf-core/viral-recon website (https://nf-co.re/viralrecon/usage), the workflow was initiated as outlined on the project's data portal (https://go.wisc.edu/4134pl). The output "variants_long_table.csv" from iVar was made into a pivot table in Microsoft Excel to make Supplemental Table 2. Because called variant frequencies differ between sequencing replicates from each time point, we decided to display the results from each replicate for the sake of transparency. Codons with variants detected in at least one sequence replicate from each time point were selected from Supplemental Table 2 and sorted by gene and frequency to make Supplemental Table 3. The presence of a particular called variant in one sequence replicate indicates that that variant could be present in the sample. The absence of a called variant in a replicate, on the other hand, does not prove its absence from the sample. Thus, we decided to include variants in Supplemental Table 3 even if they were only present in one sequence replicate for each time point.

**12s rRNA Sequencing**

12s rRNA testing of Facility Line B samples in June of 2022 was performed essentially as described in Klymus et al. 2017.[7] The primary RBD RT-PCR was performed using the Superscript IV One-Step RT-PCR System (Thermo Fisher Scientific,12594100, Waltham, MA, USA). Primary RT-PCR amplification was performed as follows: 25 °C (2:00) + 50°C (20:00) + 95°C (2:00) + [95°C (0:15) + 55°C (0:30) + 72°C (1:00)] × 30 cycles using primers 5'-acactctttccctacacgacgctcttc-cgatctACTGGGATTAGATACCCC-3' and 5'-gtgactggagttcagacgtgtgctcttccgatctTAGAACAGGCTCCTCTAG-3'. Adapter sequences were added and sequencing was performed using the same parameters as amplicon sequencing (see "Identification of cryptic lineages in wastewater with non-Omicron RT-PCR amplification and amplicon sequencing" above).
For sequencing from rRNA templates, dereplicated reads with a minimum unique count of 10 were mapped with Bowtie2 to a collected reference index of mitochondrial and rRNA related animal sequences from NCBI's nucleotide and refseq databases (https://www.ncbi.nlm.nih.gov/). Mapped rRNA sequences were reviewed for matching of specific organisms.

**Nasal swab collection and testing**

In July 2022, after high concentrations of SARS-CoV-2 were detected in the wastewater samples collected from Facility Line B, WDHS coordinated with the local public health department to offer SARS-CoV-2 testing via anterior nasal swabs to all employees of the facility. The decision to offer clinical samping to employees was made with the knowledge that at least one employee was likely harboring a SARS-CoV-2 infection with the goal of ensuring that that employees requiring COVID-19 diagnosis and/or medical services were provided with an opportunity to get tested, ask questions, and engage with the local public health department. Nasal swabs were collected by local health department staff on a designated date at the facility location. Samples were tested by RT-PCR using a clinical laboratory services contracted by the public health department. All testing was voluntary. WDHS has arranged for whole genome sequencing of any positive specimens in order to further characterize the virus, but such steps were not necessary. Further, public health actions plans were developed in collaboration with the local health department to map out potential public health follow-up in the event of a positive test, which included contingencies for isolation, additional sequencing, and additional sampling in the event that a recent or chronic SARS-CoV-2 infection were to be detected. Among approximately 30 employees, 19 were tested, and none were positive. Employees were confidentially notified of their test results, and the business manager was provided with a summary of results.

**Virus culture**

To remove debris, samples were centrifuged twice at 3,500 rpm at 4°C for 15 minutes and then passed through a 0.8 µM syringe filter (Agilent, Santa Clara, CA, USA) or left unfiltered. Samples (1ml) were incubated on nearly confluent Vero E6-TMPRSS2 (JCRB1819) or Vero E6-TMPRSS2/hACE2 cells (from Barney Graham, NIH) seeded the day prior in TC252 cm flasks for 1 hour at 370C. After the incubation, cells were washed twice and media was added back to the cells. The media contained 2-times the normal concentration of penicillin, streptomycin and amphotericin along with chloramphenicol. Cells were monitored daily for potential virus-induced cytopathic effects. After 10 days, a blind passage was performed using the entire volume of media (~4 ml) to fresh, nearly confluent cells seeded the day prior in TC1752 cm flasks.

**Variant proportion assessment**

Variant proportions were assessed from WGS data using Freyja v.1.3.11, a tool previously developed to estimate the proportions of SARS-CoV-2 variants in deep sequence data containing mixed populations (10.1038/s41586-022-05049-6).[8] Briefly, BAM files generated using viralrecon were processed by Freyja to create the variant and depth files (Wuhan-Hu-1 reference genome: MN908947.3). Variant proportions were assessed utilizing the median estimates obtained via the Freyja bootstrap *boot* function (nb = 10). The UShER barcode was updated on March 20th, 2023.

**Root-to-tip regression**

To generate Figure 4a, we first downloaded from GenBank all full consensus genomes for SARS-CoV-2 belonging to Pango lineage B.1.234 (the inferred parent of WI-CL-001) and collected from specimens in the Midwest region (Illinois, Indiana, Iowa, Kansas, Michigan, Minnesota, Missouri, Nebraska, North Dakota, Ohio, South Dakota, and Wisconsin). The accession numbers for this dataset can be found on the GitHub repository accompanying this repository. The dataset is composed of 304 individual genome sequences collected between 2020-05-04 and 2021-05-01, which represents all the available B.1.234 sequences for the Midwest region available on GenBank. The dataset was filtered to exclude incomplete and low-quality sequences and to retain no more than 50 isolates per state. The list of accession numbers for the filtered isolates can also be found on the GitHub repository accompanying this manuscript. A total of 268 sequences were ultimately aligned to the Wuhan-Hu-1 reference sequence MN908947.3 using MAFFT (v7.505).[9] A maximum likelihood phylogenetic tree was inferred using iqtree (v.2.2.0.3) with a molecular clock and distances obtained through treetime (v0.9.3).[10,11] The analysis was conducted independently for the wastewater samples (WSLH-222, WSLH-223, WSLH-230, and WSLH-231) and root-to-tip distances for all strains were visualized in R (ggplot, dplyr).[12] Phylogeny was visualized and annotated with FigTree (v.1.4.4).[13] Scripts are available in the GitHub repository accompanying this manuscript (https://github.com/tcflab/wisconsin_cryptic_lineages).

**Analyses for natural selection**

Variants obtained through the nf-core/viralrecon workflows were processed using custom Python scripts (see Data Availability) to generate panels b-d in Figure 4. The multiple replicates for each collection date were used to obtain the intersection of variants, that is, variants that were found in all replicates for each collection date. The frequencies and depth of the resulting variants were recalculated. Variants differing from reference sequence Wuhan-Hu-1 (MN908947.3) were classified as non-synonymous (Non-syn), synonymous (Syn), insertions-deletions (indels), or others (including nonsense and frameshift mutations) using SnpEff (v.5.0).[14] Synonymous and non-synonymous point mutations were quantified and compared between timepoints, and 95% confidence intervals obtained from the relative risk (RR) of every nucleotide substitution against its inverted change (i.e., RR= `A>C` / `C>A` ) using SciPy's relative_risk function (v.1.9.3).[15] To obtain the proportion of variants per site, we enumerated synonymous and non-synonymous substitutions across the SARS-CoV-2 genome, and obtained the proportion against the number of synonymous and non-synonymous sites, respectively, using SNPGenie (v.2019.10.31).[16] A binomial probability distribution was implemented to obtain the 95% confidence intervals via SciPy's binomtest function (v.1.9.3). A Mann-Whitney two-sided test was applied to test the difference between $\pi N$ and $\pi S$ on each gene, while a one-sided test was used to test for an enrichment of the $\pi N$ value of Spike against the $\pi N$ value on the other genes. To obtain synonymous and nonsynonymous divergence values (panel e), the average Hamming distance between B.1.234 isolates (dataset used in Figure 4a) and the MN908947.3 reference sequence was calculated as has been done previously for other coronaviruses.[11] Divergence was obtained over a sliding window of 36 days by dividing the observed synonymous and non-synonymous differences between the isolate and reference by the total possible number of synonymous and nonsynonymous nucleotide substitutions. Only windows that contained at least 2 sequences were considered for the analysis. Divergence values were independently calculated for each of the wastewater timepoints against the MN908947.3 reference sequence. Plot was visualized using Matplotlib.[17] Scripts are available in the GitHub repository accompanying this manuscript (https://github.com/tcflab/wisconsin_cryptic_lineages).

### Supplementary Figures

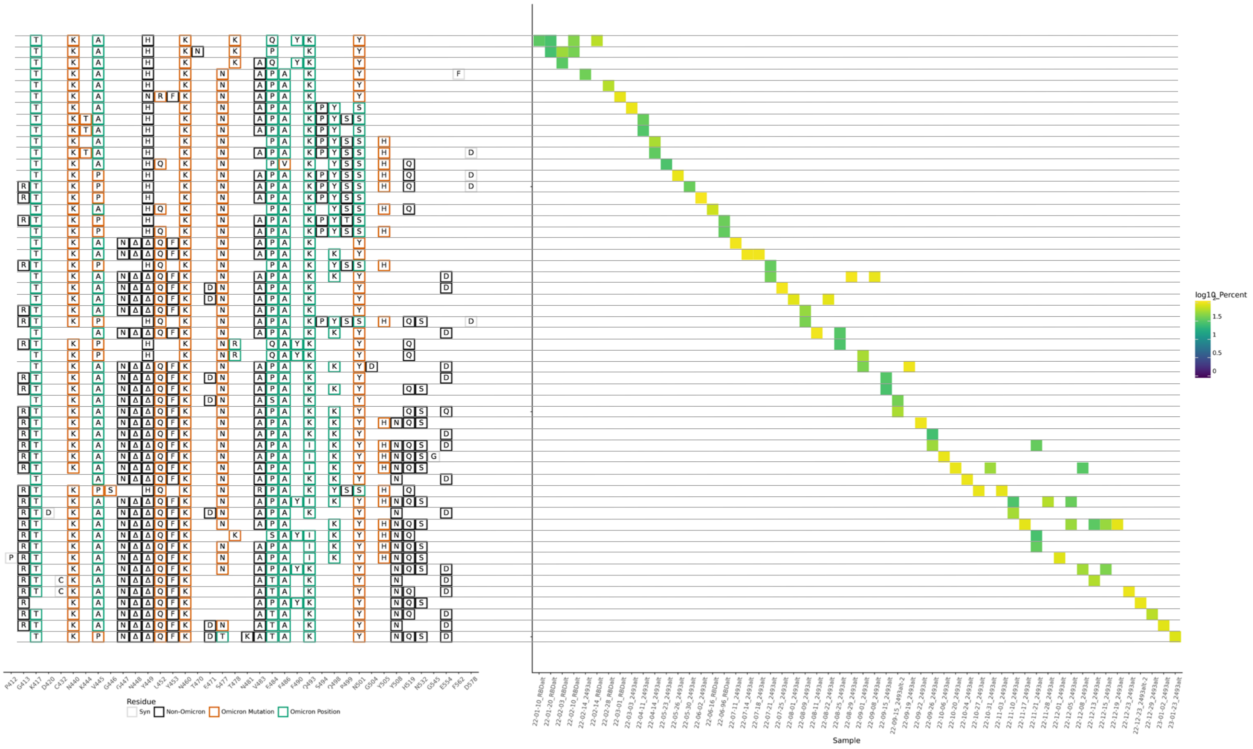

**Supplemental Figure 1.** All variants above 25% frequency in WI-CL-001 RBD sequences from Sub-District 5 are shown on the left side and are organized by haplotype. The presence and abundance within a sample are shown on the right side.

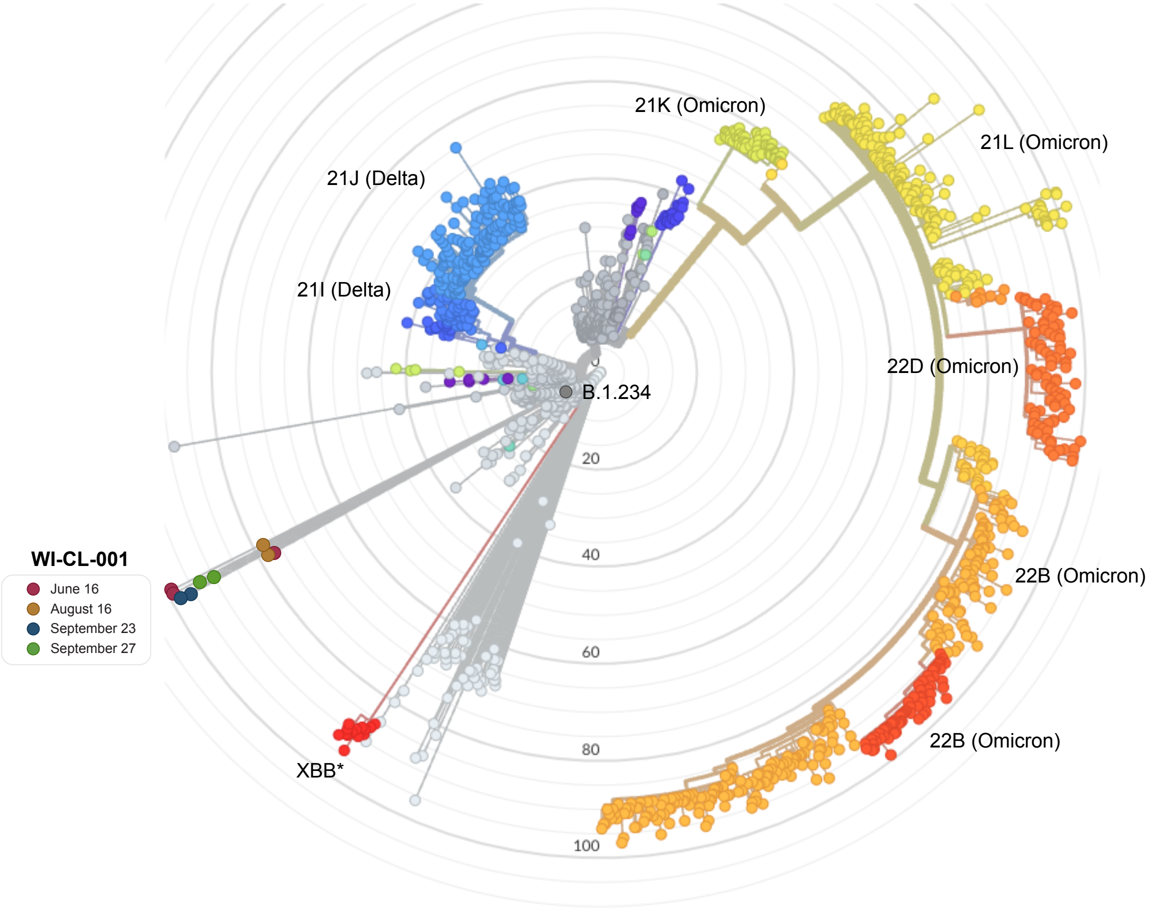

**Supplemental Figure 2.** Radial phylogenetic tree generated by Nextclade.[1] Consensus fasta files generated for each sequence replicate of Facility Line B samples are shown. Although differences exist between some replicate sequences, WI-CL-001 greatly diverges from its B.1.234 backbone and is similarly divergent to Omicron lineages.

#### Supplementary Tables

| Species | Common Name | Facility Line B (June) | |
|---|---|---|---|
| | | Average Count | Average Abundance |
| Homo sapiens | Human | 116,842.68 | 88.22% |
| Unmatched | Unmatched | 12,522.80 | 9.46% |
| Pan paniscus | Human, chimp, but more likely human | 4,507.00 | 3.40% |
| Poor match | | 794.00 | 0.60% |
| Elephantulus fuscipes | Human - mismatch, human | 72.00 | 0.05% |
| Gallus gallus | Bird, Chicken | 49.00 | 0.04% |
| Bos taurus | Cattle | 30.00 | 0.02% |
| Pan troglodytes | Human, chimp, but also human | 29.00 | 0.02% |
| Rhynchocyon petersi | Human - mismatch, human | 27.00 | 0.02% |
| Catopuma temminckii | Human - Asian golden cat, but also human | 24.66 | 0.02% |
| Oreochromis niloticus | Fish, Tilapia | 14.00 | 0.01% |

**Supplemental Table 1.** Abundance of contributing species to the June Facility Line B sample. 12S ribosomal RNA (rRNA) sequencing was performed on two replicates of the June sample of Facility Line B. The average count and abundance between these two samples is shown. *Homo sapiens* (human) was found to be the predominant contributor to this sample. Cow and chicken rRNA were the second and third most abundant (<1% each).

**Supplemental Table 2 (Appendix 2).** All SARS-CoV-2 variants identified in Facility Line B whole genome sequences. Illumina whole genome sequencing data of overlapping PCR amplicons from Facility Line B were processed using the nf-core/viralrecon workflow. Variants from reference Wuhan-Hu-1 called by iVar at a frequency of at least 25% in any of the sequencing replicates from any of our four sampling time points are shown along with their location in the genome and predicted protein impact. The content of this table has not been altered in any way from the content of the iVar "variants_long_table.csv". In some cases there are inaccuracies with the variant calls that we have left for the sake of reproducibility. See the Supplemental Table 3 legend for a specific example of this. See the excel file named "Appendix 2".

| Variant frequency summed by codon | | 16-Jun-22 3 replicates | | | 16-Aug-22 2 replicates | | 23-Sep-22 2 replicates | | 27-Sep-22 2 replicates | | | | | |
|---|---|---|---|---|---|---|---|---|---|---|---|---|---|---|
| **Gene** | **Codon** | **#1 WSLH-152** | **#2 WSLH-229** | **#3 WSLH-231** | **#1 WSLH-206** | **#2 WSLH-230** | **#1 WSLH-221** | **#2 WSLH-222** | **#1 WSLH-223** | **#2 WSLH-224** | **Sum** | **Average** | **Variants** | **B.1.234** |
| ORF1ab | L4715 | 1.28 | 1.24 | 1.17 | 1.3 | 1.23 | 1.24 | 1.35 | 1.2 | 1.31 | 11.32 | 1.26 | L4715_V4717del; L4715L | * |
| ORF1ab | V38 | 1 | 1 | 1 | 1 | 1 | 1 | 1 | 1 | 1 | 9 | 1.00 | V38A | |
| ORF1ab | F924 | 1 | 1 | 1 | 1 | 1 | 1 | 1 | 1 | 1 | 9 | 1.00 | F924F | |
| ORF1ab | A1234 | 1 | 1 | 1 | 1 | 1 | 1 | 1 | 1 | 1 | 9 | 1.00 | A1234A | |
| ORF1ab | D2980 | 1 | 1 | 1 | 1 | 1 | 1 | 1 | 1 | 1 | 9 | 1.00 | D2980G | * |
| ORF1ab | K1795 | 1 | 1 | 1 | 1 | 1 | 1 | 1 | 0.99 | 1 | 8.99 | 1.00 | K1795Q | |
| ORF1ab | *6668Wext*? | 1 | 0.99 | 1 | 1 | 1 | 1 | 1 | 1 | 1 | 8.99 | 1.00 | stop_lost | |
| ORF1ab | T4461 | 1 | 0.99 | 1 | 1 | 1 | 0.99 | 1 | 1 | 1 | 8.98 | 1.00 | T4461I | |
| ORF1ab | L3606 | 0.99 | 1 | 1 | 0.99 | 0.99 | 1 | 1 | 1 | 1 | 8.97 | 1.00 | L3606V | |
| ORF1ab | S2625 | 1 | 1 | 1 | 1 | 1 | 0.99 | 0.99 | 0.99 | 0.99 | 8.96 | 1.00 | S2625S | |
| ORF1ab | V665 | 1 | 1 | 1 | 0.99 | 1 | 1 | 1 | 0.96 | 1 | 8.95 | 0.99 | V665I | * |
| ORF1ab | A2199 | 1 | 0.98 | 0.98 | 1 | 0.98 | 1 | 1 | 0.95 | 1 | 8.89 | 0.99 | A2199T | |
| ORF1ab | R24 | 0.94 | 0.99 | 0.87 | 0.98 | 0.99 | 0.93 | 0.92 | 0.89 | 0.93 | 8.44 | 0.94 | R24C | |
| ORF1ab | L6710 | 0.9 | 1 | 0.68 | 0.96 | 1 | 1 | 0.95 | 0.92 | 1 | 8.41 | 0.93 | L6710L | |
| ORF1ab | L6714 | 0.9 | 1 | 0.68 | 0.96 | 1 | 1 | 0.95 | 0.91 | 1 | 8.4 | 0.93 | L6714L | |
| ORF1ab | S3149 | 0.94 | 0.46 | 1 | 0.98 | 1 | 0.98 | 0.95 | 0.9 | 1 | 8.21 | 0.91 | S3149F | |
| ORF1ab | I3148 | 0.93 | 0.41 | 0.92 | 0.98 | 0.91 | 0.98 | 0.95 | 0.9 | 1 | 7.98 | 0.89 | I3148L; I3148P | |
| ORF1ab | L123 | 0.78 | 0.87 | 0.96 | 0.89 | 0.57 | 0.86 | 0.92 | 0.96 | 0.97 | 7.78 | 0.86 | L123F | |
| ORF1ab | F3677 | 0.65 | 0.86 | 0.98 | 0.79 | 0.53 | 1 | 0.87 | 0.92 | 0.84 | 7.44 | 0.83 | F3677I | |
| ORF1ab | T3258 | 0.93 | 0.99 | | 0.94 | 0.99 | 0.79 | 0.94 | 0.91 | 0.88 | 7.37 | 0.92 | T3258T | |
| ORF1ab | D4719 | 0.77 | 0.8 | 0.8 | 0.77 | 0.7 | 0.85 | 0.84 | 0.92 | 0.87 | 7.32 | 0.81 | D4719fs | |
| ORF1ab | F1779 | 0.49 | 0.52 | 0.81 | 0.92 | 1 | 0.91 | 0.95 | 0.77 | 0.66 | 7.03 | 0.78 | F1779L; F1779L | |
| ORF1ab | T3058 | 0.53 | 0.52 | 1 | 0.75 | 1 | 0.84 | 0.75 | 0.79 | 0.84 | 7.02 | 0.78 | T3058V; T3058A |  |
| ORF1ab | K6473 | 0.85 | 0.98 | 0.61 | 0.82 | 0.67 | 0.97 | 0.8 | 0.62 | 0.65 | 6.97 | 0.77 | K6473K | |
| ORF1ab | L92 | 0.67 | 0.74 | 0.63 | 0.67 | 0.81 | 0.82 | 0.8 | 0.87 | 0.83 | 6.84 | 0.76 | L92F | |
| ORF1ab | C2012 | 0.69 | 0.64 | 1 | 0.71 | 0.76 | 0.7 | 0.75 | 0.76 | 0.82 | 6.83 | 0.76 | C2012S | |
| ORF1ab | S167 | 0.75 | 1 | 0.54 | 0.68 | 0.81 | 0.77 | 0.67 | 0.55 | 0.84 | 6.61 | 0.73 | S167G | |
| ORF1ab | M85 | 0.65 | 0.74 | 0.52 | 0.63 | 0.73 | 0.75 | 0.75 | 0.84 | 0.82 | 6.43 | 0.71 | M85fs | |
| ORF1ab | N9 | 0.58 | 0.45 | 0.46 | 0.69 | 0.71 | 0.91 | 0.86 | 0.87 | 0.81 | 6.34 | 0.70 | N9N | |
| ORF1ab | C2009 | 0.6 | 0.56 | 0.72 | 0.69 | 0.63 | 0.7 | 0.74 | 0.76 | 0.83 | 6.23 | 0.69 | C2009G; C2009V | |
| ORF1ab | R6968 | 0.77 | 1 | 0.76 | 0.56 | 0.82 | 0.57 | 0.65 | 0.59 | 0.5 | 6.22 | 0.69 | R6968K | |
| ORF1ab | R5036 | 0.68 | 0.71 | 0.58 | 0.65 | 0.58 | 0.81 | 0.68 | 0.78 | 0.7 | 6.17 | 0.69 | R5036H | |
| ORF1ab | S4646 | 0.66 | 0.53 | 0.79 | 0.59 | 0.45 | 0.75 | 0.65 | 0.83 | 0.81 | 6.06 | 0.67 | S4646C | |
| ORF1ab | K2059 | 0.69 | 0.61 | 0.76 | 0.72 | 0.47 | 0.62 | 0.62 | 0.77 | 0.79 | 6.05 | 0.67 | K2059R | * |
| ORF1ab | R6997 | 0.76 | 1 | 0.77 | 0.6 | 0.72 | 0.49 | 0.43 | 0.5 | 0.77 | 6.04 | 0.67 | R6997P | |
| ORF1ab | T4652 | 0.59 | 0.62 | 0.59 | 0.65 | 0.51 | 0.7 | 0.69 | 0.82 | 0.8 | 5.97 | 0.66 | T4652S | |
| ORF1ab | S1188 | | 0.97 | | 0.57 | 0.93 | 0.65 | 0.96 | 0.88 | 0.95 | 5.91 | 0.84 | S1188L | |
| ORF1ab | A1306 | 0.58 | 0.62 | 0.78 | 0.61 | 0.53 | 0.5 | 0.48 | 0.58 | 0.8 | 5.48 | 0.61 | A1306T | |
| ORF1ab | G5231 | 0.62 | 0.49 | 0.57 | 0.55 | 0.61 | 0.53 | 0.58 | 0.56 | 0.53 | 5.04 | 0.56 | G5231R | |
| ORF1ab | K5057 | 0.44 | 0.49 | 0.28 | 0.56 | 0.47 | 0.79 | 0.63 | 0.74 | 0.57 | 4.97 | 0.55 | K5057N | |
| ORF1ab | T727 | 0.31 | 0.7 | 0.29 | 0.65 | 0.84 | 0.38 | 0.52 | 0.59 | 0.67 | 4.95 | 0.55 | T727I | |
| ORF1ab | K1988 | 0.43 | 0.4 | 0.45 | 0.5 | 0.42 | 0.59 | 0.67 | 0.7 | 0.73 | 4.89 | 0.54 | K1988E | |
| ORF1ab | R43 | 0.47 | 0.91 | 0.84 | 0.35 | 0.48 | 0.44 | 0.59 | 0.36 | 0.36 | 4.8 | 0.53 | R43R | |
| ORF1ab | P4638 | 0.46 | 0.32 | 0.51 | 0.47 | 0.44 | 0.63 | 0.56 | 0.64 | 0.68 | 4.71 | 0.52 | P4638fs | |
| ORF1ab | E2070 | 0.47 | 0.31 | 0.48 | 0.57 | 0.33 | 0.54 | 0.5 | 0.66 | 0.6 | 4.46 | 0.50 | E2070G | |
| ORF1ab | T2823 | 0.54 | | 0.44 | 0.58 | 0.35 | 0.6 | 0.76 | 0.57 | 0.55 | 4.39 | 0.55 | T2823A | |
| ORF1ab | N128 | | | 0.42 | 0.44 | 0.31 | 0.78 | 0.68 | 0.92 | 0.83 | 4.38 | 0.63 | N128N | |
| ORF1ab | G82 | 0.4 | 0.47 | 0.48 | 0.36 | 0.57 | 0.47 | 0.48 | 0.6 | 0.52 | 4.35 | 0.48 | G82fs | |
| ORF1ab | I2385 | 0.35 | | | 0.55 | 0.49 | 0.68 | 0.71 | 0.78 | 0.74 | 4.3 | 0.61 | I2385I | |
| ORF1ab | N854 | 0.49 | 0.45 | 0.39 | 0.49 | 0.61 | 0.49 | 0.62 | 0.27 | 0.35 | 4.16 | 0.46 | N854D | |
| ORF1ab | M4555 | 0.45 | 0.37 | 0.68 | 0.34 | 0.46 | 0.49 | 0.28 | 0.49 | 0.57 | 4.13 | 0.46 | M4555fs | |
| ORF1ab | I1159 | 0.48 | 0.4 | 1 | 0.45 | 0.34 | 0.45 | | 0.34 | 0.43 | 3.89 | 0.49 | I1159T | |
| ORF1ab | M4521 | 0.32 | 0.26 | 0.26 | 0.31 | 0.28 | 0.62 | 0.68 | 0.67 | 0.47 | 3.87 | 0.43 | M4521fs | |
| ORF1ab | G445 | 0.34 | 0.43 | 0.38 | 0.42 | 0.58 | 0.52 | 0.58 | 0.28 | 0.29 | 3.82 | 0.42 | G445C | |
| ORF1ab | N2006 | 0.32 | | | 0.47 | 0.46 | 0.58 | 0.63 | 0.62 | 0.73 | 3.81 | 0.54 | N2006N | |
| ORF1ab | H1160 | 0.37 | 0.42 | | 0.31 | 0.49 | 0.55 | 0.82 | 0.44 | 0.39 | 3.79 | 0.47 | H1160Y; H1160Q | |
| ORF1ab | L5028 | 0.38 | 0.56 | 0.46 | 0.34 | 0.37 | 0.37 | 0.34 | 0.53 | 0.42 | 3.77 | 0.42 | L5028F | |
| ORF1ab | S4655 | 0.26 | 0.45 | | 0.35 | | 0.65 | 0.6 | 0.68 | 0.65 | 3.64 | 0.52 | S4655I | |

| | | | | | | | | | | | | | | |
|---|---|---|---|---|---|---|---|---|---|---|---|---|---|---|
| ORF1ab | S74 | | 0.38 | 0.45 | 0.28 | 0.48 | 0.45 | 0.52 | 0.55 | 0.47 | 3.58 | 0.45 | S74T | |
| ORF1ab | E5023 | 0.37 | 0.43 | 0.4 | 0.31 | 0.34 | 0.37 | 0.31 | 0.46 | 0.39 | 3.38 | 0.38 | E5023_L5024del | |
| ORF1ab | Q22 | 0.27 | | 0.46 | 0.36 | 0.41 | 0.41 | 0.45 | 0.51 | 0.41 | 3.28 | 0.41 | Q22K | |
| ORF1ab | E57 | 0.38 | 0.62 | 0.4 | 0.29 | 0.27 | 0.33 | 0.44 | | 0.27 | 3 | 0.38 | E57K | |
| ORF1ab | L27 | | | 0.53 | 0.33 | 0.36 | 0.41 | 0.45 | 0.51 | 0.41 | 3 | 0.43 | L27F | |
| ORF1ab | S4740 | | 0.64 | | 0.25 | | 0.47 | 0.47 | 0.43 | 0.31 | 2.57 | 0.43 | S4740P | |
| ORF1ab | A2584 | 0.3 | | | 0.32 | 0.8 | | 0.52 | 0.27 | | 2.21 | 0.44 | A2584T | |
| ORF1ab | T4738 | 0.28 | | | | 0.25 | 0.39 | 0.34 | 0.4 | 0.43 | 2.09 | 0.35 | T4738T | |
| ORF1ab | H165 | 0.25 | | 0.46 | 0.3 | | | 0.33 | 0.45 | | 1.79 | 0.36 | H165Y | |
| ORF1ab | S4986 | | 0.38 | | 0.26 | | 0.35 | 0.45 | 0.29 | | 1.73 | 0.35 | S4986F | |
| S | N460 | 1 | 1 | 1 | 1 | 1 | 1 | 1 | 1 | 1 | 9 | 1.00 | N460K | |
| S | Q493 | 1 | 1 | 1 | 1 | 1 | 1 | 1 | 1 | 1 | 9 | 1.00 | Q493K | |
| S | D614 | 1 | 1 | 1 | 1 | 1 | 1 | 1 | 1 | 1 | 9 | 1.00 | D614G | * |
| S | D1153 | 1 | 1 | 1 | 1 | 1 | 1 | 1 | 1 | 1 | 9 | 1.00 | D1153A | |
| S | T1231 | 1 | 1 | 0.99 | 1 | 1 | 1 | 1 | 1 | 1 | 8.99 | 1.00 | T1231I | |
| S | K417 | 0.98 | 1 | 1 | 1 | 1 | 1 | 1 | 1 | 1 | 8.98 | 1.00 | K417T | |
| S | Y449 | 0.98 | 1 | 1 | 1 | 1 | 1 | 1 | 1 | 1 | 8.98 | 1.00 | Y449H | |
| S | V1176 | 1 | 1 | 0.99 | 1 | 0.99 | 1 | 1 | 1 | 1 | 8.98 | 1.00 | V1176F | |
| S | S940 | 0.98 | 1 | 0.99 | 1 | 1 | 1 | 1 | 1 | 1 | 8.97 | 1.00 | S940T | |
| S | D215 | 1 | 1 | 0.98 | 1 | 0.99 | 1 | 1 | 1 | 1 | 8.97 | 1.00 | D215G | |
| S | F486 | 0.99 | 1 | 1 | 0.99 | 1 | 1 | 1 | 0.98 | 1 | 8.96 | 1.00 | F486A | |
| S | R214 | 1 | 0.98 | 0.99 | 1 | 0.99 | 0.99 | 0.99 | 1 | 1 | 8.94 | 0.99 | R214S | |
| S | Q14 | 0.99 | 1 | 1 | 0.99 | 0.99 | 0.95 | 1 | 1 | 1 | 8.92 | 0.99 | Q14L | |
| S | R1185 | 0.99 | 0.99 | 0.99 | 1 | 0.99 | 0.99 | 0.99 | 0.99 | 0.99 | 8.92 | 0.99 | R1185H | |
| S | K150 | 0.98 | 0.87 | 1 | 0.99 | 1 | 1 | 0.98 | 1 | 1 | 8.82 | 0.98 | K150N | |
| S | V367 | 0.8 | 1 | 1 | 0.97 | 1 | 1 | 1 | 1 | 1 | 8.77 | 0.97 | V367F | |
| S | L828 | 0.99 | 0.99 | 0.99 | 0.99 | 1 | 0.91 | 0.79 | 0.99 | 1 | 8.65 | 0.96 | L828V; L828F | |
| S | S375 | 0.84 | 0.95 | 0.99 | 0.91 | 1 | 0.98 | 0.98 | 1 | 0.99 | 8.64 | 0.96 | S375P | |
| S | P384 | 0.99 | 0.74 | 0.89 | 0.99 | 0.99 | 1 | 1 | 1 | 1 | 8.6 | 0.96 | P384L | |
| S | Q498 | 1.88 | 1.96 | 1.98 | 0.32 | 0.38 | 0.65 | 0.62 | 0.37 | 0.43 | 8.59 | 0.95 | Q498*; Q498R; Q498R; Q498H | |
| S | S477 | 0.98 | 1 | 1 | 0.98 | 0.98 | 0.91 | 0.91 | 0.81 | 1 | 8.57 | 0.95 | S477N | |
| S | E484 | 1 | 1 | 1 | 0.98 | 1 | 0.95 | 0.91 | 0.79 | 0.84 | 8.47 | 0.94 | E484P; E484Q | |
| S | T19 | 0.86 | 0.86 | 0.85 | 0.94 | 0.98 | 0.87 | 1 | 1 | 1 | 8.36 | 0.93 | T19K | |
| S | N501 | 0.93 | 0.98 | 0.99 | 0.82 | 0.88 | 0.89 | 0.9 | 0.98 | 0.97 | 8.34 | 0.93 | N501Y; N501S | |
| S | V445 | 0.74 | 0.62 | 0.85 | 1 | 0.99 | 1 | 0.99 | 1 | 1 | 8.19 | 0.91 | V445P; V445A | |
| S | N137 | 0.91 | 0.89 | 0.86 | 0.98 | 0.87 | 0.82 | 0.78 | 0.96 | 0.89 | 7.96 | 0.88 | N137K | |
| S | R346 | 0.74 | 1 | 1 | 0.95 | 0.39 | 1 | 1 | 0.79 | 1 | 7.87 | 0.87 | R346T | |
| S | S1261 | 0.84 | 1 | 0.92 | 0.77 | 0.79 | 0.9 | 0.7 | 0.97 | 0.96 | 7.85 | 0.87 | S1261F | |
| S | D1260 | 0.83 | 0.95 | 0.82 | 0.69 | 0.88 | 0.64 | 0.54 | 0.86 | 0.75 | 6.96 | 0.77 | D1260E | |
| S | A260 | 0.99 | 0.98 | 0.38 | 0.94 | 0.54 | 0.46 | 0.86 | | 0.99 | 6.14 | 0.77 | A260V | |
| S | H69 | 0.58 | 0.91 | 0.69 | 0.57 | 0.83 | 0.6 | 0.58 | 0.56 | 0.61 | 5.93 | 0.66 | H69_V70del | |
| S | R408 | 0.78 | | 0.84 | 0.77 | 0.48 | 0.85 | 0.8 | 0.62 | 0.58 | 5.72 | 0.72 | R408K | |
| S | H66 | 0.38 | 1 | 0.35 | 0.52 | 0.65 | 0.96 | 0.79 | 0.73 | 0.31 | 5.69 | 0.63 | H66R | |
| S | N440 | 0.94 | 0.98 | 0.85 | 0.43 | 0.3 | 0.26 | 0.39 | 0.55 | 0.77 | 5.47 | 0.61 | N440K | |
| S | Q173 | 0.56 | 0.48 | 1 | 0.41 | 0.35 | 0.82 | 0.7 | 0.55 | 0.33 | 5.2 | 0.58 | Q173K; Q173R; Q173R | |
| S | H519 | 0.67 | | 0.66 | 0.38 | 0.68 | 0.62 | 0.85 | 0.72 | 0.57 | 5.15 | 0.64 | H519Q | |
| S | Y248 | | 0.98 | 0.98 | | 0.98 | | 0.99 | | 1 | 4.93 | 0.99 | Y248R; Y248H | |
| S | F1052 | 0.32 | 1 | | 0.47 | | 0.82 | 0.91 | 0.54 | 0.76 | 4.82 | 0.69 | F1052L | |
| S | L24 | 0.59 | | | 0.61 | 0.69 | 0.66 | 0.95 | 0.68 | 0.42 | 4.6 | 0.66 | L24S | |
| S | F374 | | 0.92 | | 0.36 | 0.86 | 0.99 | 0.35 | 0.39 | 0.45 | 4.32 | 0.62 | F374Y | |
| S | S373 | 0.42 | | | 0.71 | 0.29 | 0.89 | 0.84 | 0.5 | 0.54 | 4.19 | 0.60 | S373fs | |
| S | L48 | 0.44 | | | 0.55 | 0.56 | 0.78 | 0.83 | 0.7 | 0.3 | 4.16 | 0.59 | L48I | |
| S | Y145 | 0.31 | | | 0.59 | 0.63 | 0.82 | 0.77 | 0.46 | 0.43 | 4.01 | 0.57 | Y145H | |
| S | P793 | | 0.68 | | 0.44 | 0.32 | 0.77 | 0.6 | 0.41 | 0.36 | 3.58 | 0.51 | P793S | |
| S | T76 | 0.63 | | 0.56 | 0.49 | 0.38 | | 0.26 | 0.31 | 0.79 | 3.42 | 0.49 | T76I | |
| ORF3a | N152 | 1 | 1 | 1 | 1 | 1 | 1 | 1 | 1 | 1 | 9 | 1.00 | N152H | |
| ORF3a | H182 | 1 | 1 | 1 | 1 | 1 | 1 | 1 | 1 | 1 | 9 | 1.00 | H182D | |
| ORF3a | G196 | 1 | 1 | 1 | 1 | 1 | 1 | 1 | 1 | 1 | 9 | 1.00 | G196E | |
| ORF3a | C200 | 1 | 1 | 1 | 1 | 1 | 1 | 1 | 1 | 1 | 9 | 1.00 | C200Y | |
| ORF3a | L219 | | | 1 | 1 | | 1 | 0.96 | 1 | 1 | 5.96 | 0.99 | L219V | |
| ORF3a | I10 | 0.52 | 0.29 | 0.67 | 0.56 | 0.48 | | 0.4 | 0.58 | 0.55 | 4.05 | 0.51 | I10L | |

| | | | | | | | | | | | | | | |
|---|---|---|---|---|---|---|---|---|---|---|---|---|---|---|
| ORF3a | I7 | 0.44 | 0.7 | | 0.34 | 0.52 | 0.49 | 0.39 | 0.3 | | 3.18 | 0.45 | I7L | |
| M | A2 | 0.99 | 1 | 1 | 1 | 1 | 1 | 1 | 1 | 1 | 8.99 | 1.00 | A2E | |
| M | G6 | 0.99 | 0.99 | 1 | 1 | 1 | 1 | 1 | 0.99 | 1 | 8.97 | 1.00 | G6C | |
| M | L17 | 1 | 0.99 | 1 | 1 | 1 | 1 | 1 | 0.97 | 1 | 8.96 | 1.00 | L17V | |
| M | I8 | 0.96 | 0.91 | 0.8 | 0.96 | 0.94 | 0.96 | 0.92 | 0.96 | 0.91 | 8.32 | 0.92 | I8delinsSNNSEF | |
| M | V23 | 0.38 | 0.84 | 0.6 | 0.75 | 1 | 0.72 | 0.89 | 0.84 | 0.53 | 6.55 | 0.73 | V23A | |
| ORF6 | N28 | 0.99 | 1 | 0.86 | 1 | 1 | 1 | 1 | 1 | 1 | 8.85 | 0.98 | N28K | |
| ORF7a | S81 | 0.97 | 1 | 0.74 | 1 | 1 | 1 | 1 | 1 | 1 | 8.71 | 0.97 | S81P | |
| ORF7a | F59 | 0.68 | | 0.59 | 0.89 | 0.85 | 0.91 | 0.96 | 1 | 1 | 6.88 | 0.86 | F59I | |
| ORF8 | G66 | 0.31 | | 0.56 | 0.52 | 0.87 | 0.5 | 0.62 | 0.52 | 1 | 4.9 | 0.61 | G66_K68delinsE | |
| N | P326 | 0.99 | 1 | 0.99 | 1 | 1 | 1 | 1 | 0.99 | 1 | 8.97 | 1.00 | P326S | |
| N | S194 | 1 | 1 | 1 | 1 | 1 | 1 | 0.91 | 1 | 1 | 8.91 | 0.99 | S194L | * |
| N | S37 | 1 | 1 | 1 | 1 | 1 | 0.45 | 0.89 | 0.55 | 0.65 | 7.54 | 0.84 | S37P | |
| N | T16 | 1 | 0.98 | 0.99 | 1 | 0.99 | 0.4 | 0.87 | 0.43 | 0.56 | 7.22 | 0.80 | T16T | |
| N | E31 | 0.41 | 0.79 | 0.99 | 0.73 | 0.77 | 1.01 | 0.72 | 0.81 | 0.84 | 7.07 | 0.79 | E31_S33del | |
| N | Q160 | 0.59 | 0.37 | 0.84 | 0.86 | 0.54 | 0.9 | 0.7 | 1 | 0.91 | 6.71 | 0.75 | Q160Q | |
| N | S413 | 0.39 | 0.53 | | 0.57 | 0.53 | 0.55 | 0.5 | 0.54 | 0.48 | 4.09 | 0.51 | S413R | |

**Supplemental Table 3.** Shared SARS-CoV-2 variants found in June, August, and September 2022 Facility Line B sequences. Variants called by iVar[2] found in whole genome sequences at every time point in Supplemental Table 2 are shown here grouped by codon and sorted by gene and frequency. The frequencies of multiple variants within one codon are summed, leading to some codons with multiple variants having frequencies above 1.0. Variants characteristic of Pango lineage B.1.234, as noted on outbreak.info (https://outbreak.info/), are marked with an asterisk.[3] One of the variants typical of B.1.234 is P4715L. iVar called a synonymous "L4715L" variant at this position in all Facility Line B sequences, while a manual inspection of consensus sequences aligned with the Wuhan-Hu-1 reference instead found the P4715L non-synonymous variant known in B.1.234. We believe that this mistake by iVar could be due to multiple deletions in this area of the genome. We have reported it as-called for the sake of reproducibility. Due to the number of deletions and frameshifts present in our whole genome sequences, there may be more cases like this elsewhere.
